# The upper respiratory tract microbiome of indigenous Orang Asli in north-eastern Peninsular Malaysia

**DOI:** 10.1101/2020.06.02.20120444

**Authors:** D. W. Cleary, D. E. Morris, R. A. Anderson, J. Jones, A. G. Alattraqchi, N. I. A. Rahman, S. Ismail, M. S. Razali, Amin R. Mohd, Aziz A. Abd, N. K. Esa, S. Amiruddin, C. H. Chew, Simin M. H. Amat, R. Abdullah, C. C. Yeo, S. C. Clarke

**Affiliations:** Faculty of Medicine and Institute for Life Sciences, University of Southampton, Southampton, UK; Southampton NIHR Biomedical Research Centre, University Hospital Southampton NHS Trust, Southampton, UK; Faculty of Medicine, Universiti Sultan Zainal Abidin, Medical Campus, 20400 Kuala Terengganu, Terengganu, Malaysia; Faculty of Health Sciences, Universiti Sultan Zainal Abidin, Gong Badak Campus, 21300 Kuala Nerus, Terengganu, Malaysia; Faculty of Applied Social Sciences, Universiti Sultan Zainal Abidin, Gong Badak Campus, 21300 Kuala Nerus, Terengganu, Malaysia; Global Health Research Institute, University of Southampton, Southampton, UK; School of Postgraduate Studies, International Medical University, Kuala Lumpur, Malaysia; Centre for Translational Research, IMU Institute for Research, Development and Innovation (IRDI), Kuala Lumpur, Malaysia

**Keywords:** 16S rRNA, microbiome, upper respiratory tract, antimicrobial resistance, Malaysia, *Streptococcus pneumoniae*, *Haemophilus influenzae*, *Staphylococcus aureus*, *Moraxella catarrhalis*, Orang Asli, indigenous people

## Abstract

**Background:** Microbiome research has focused on populations that are predominantly of European descent, and from narrow demographics that do not capture the socio-economic and lifestyle differences which impact human health. This limits our understanding of human-host microbiota interactions in their broadest sense. Here we examined the airway microbiology of the Orang Asli, the indigenous peoples of Malaysia. In addition to exploring the carriage and antimicrobial resistance of important respiratory pathobionts, we also present the first investigation of the nasal microbiomes of these indigenous peoples, in addition to their oral microbiomes.

**Results:** A total of 130 participants were recruited to the study from Kampung Sungai Pergam and Kampung Berua, both sites in the north-eastern state of Terengganu in Peninsular Malaysia. High levels of *Staphylococcus aureus* carriage were observed, particularly in the 18-65 age group (n=17/36; 47.2% 95%CI: 30.9-63.5). The highest carriage of pneumococci was in the <5 and 5 to 17 year olds, with 57.1% (4/7) and 49.2% (30/61) respectively. Sixteen pneumococcal serotypes were identified, the most common being the non-vaccine type 23A (14.6%) and the vaccine type 6B (9.8%). The nasal microbiome was significantly more diverse in those aged 5-17 years compared to 50+ years (p = 0.023). In addition, samples clustered by age (PERMANOVA analysis of the Bray-Curtis distance, *p* = 0.001). Hierarchical clustering of Bray-Curtis dissimilarity scores revealed six microbiome types. The largest cluster (n=28; 35.4%) had a marked abundance of *Corynebacterium*.

Others comprised *Corynebacterium* with *Dolosigranulum*, two clusters were definable by the presence of *Moraxella*, one with and the other without *Haemophilus*, a small grouping of *Delftia/ Ochrobactum* profiles and one with *Streptococcus*. No *Staphylococcus* profiles were observed. In the oral microbiomes *Streptococcus, Neisseria* and *Haemophilus* were dominant. Lower levels of *Prevotella, Rothia, Porphyromonas, Veillonella* and *Aggregatibacter* were also among the eight most observed genera.

**Conclusions:** We present the first study of Orang Asli airway microbiomes and pathobiont microbiology. Key findings include the prevalence of pneumococcal serotypes that would be covered by pneumococcal conjugate vaccines if introduced into a Malaysian national immunisation schedule, and the high level of *S. aureus* carriage. The dominance of *Corynebacterium* in the airway microbiomes is particularly intriguing given its’ consideration as a potentially protective commensal with respect to acute infection and respiratory health.

## Background

Much microbiome research has, to-date, been focused on populations that are predominantly of European descent, and from demographics that do not capture the socio-economic and lifestyle challenges which impact human health [1]. The study of non-Western, unindustrialised populations is therefore an important extension of microbiome research so that we may understand human-host microbiota interactions in their broadest sense. Notable endeavours here, which have principally focussed on gut microbiomes, include studies of the Hadza hunter-gatherers of Tanzania [2], Amerindians of South America [3, 4], agriculturalist communities in Burkina Faso [5] Malawi and Venezuela [6] and the Cheyenne and Arapaho Tribes of North America [7]. Fewer studies have gone beyond gut microbiomes to sample additional body sites, such as those of the airways. The analysis of salivary microbiota of the Batwa Pygmies [8], and the Yanomami of the Venezuelan Amazon [3] have shown the benefits of including these body sites where, respectively, previously undiscovered genera have been identified and novel combinations of taxa described. However, there remains a paucity of data regarding microbiomes of other anatomical sites of the upper respiratory tract (URT). With respiratory infectious diseases continuing to be a significant component of global morbidity and mortality [9], the interest in the microbiome of the upper airways stems from its’ importance in an individual’s susceptibility to respiratory infection, in part through the presence of resilient taxa which prevent colonisation and/or outgrowth of specific pathobionts [10]. Understanding the variability in airway microbiomes is a key first step to leveraging these interactions for health. To date no study has undertaken a comprehensive analysis of the URT microbiome of the indigenous populations of Peninsular Malaysia, the Orang Asli.

Although the name Orang Asli was only introduced around 1960 by the British, as an ethnic group they are believed to be the first settlers of Peninsular Malaysia, moving from Northern Thailand, Burma and Cambodia 3-8000 years ago. Orang Asli is in fact a catch-all term for three tribal groups, the Senoi, Proto-Malays and Negrito, each of which in turn can be further divided into six ethnic groups [11]. They number ~179 000, 0.6% of the population of Malaysia but are disproportionately impacted by economic marginalisation and discrimination [12]. As a consequence, nearly 80% of the Orang Asli population are beneath the poverty line compared to 1.4% of the national population, and this hardship is reflected in a 20-year lower average life expectancy of just 53 years [13]. Several studies have highlighted the susceptibility of these populations to specific diseases [14, 15]. More recently a study of 73 adults from the semi-urbanized Temiar, an Orang Asli tribe of Kampong Pos Piah, in Perak, examined salivary microbiomes in the context of obesity – a growing concern and evidence of the epidemiological shifts in disease burden as these communities leave their more traditional existences [16].

Here we examined the microbiology of the upper airway of two Orang Asli communities located in Terengganu state, North-east of Peninsular Malaysia. The nasopharyngeal and nasal (anterior nares) carriage of important pathobionts was determined by culture, and the resistance of these isolates to clinically relevant antibiotics tested. We also present the first investigation of the nasal microbiomes of these indigenous peoples, in addition to their oral microbiomes.

## Results

A total of 130 participants were recruited to the study, with 68 from Kampung Sungai Pergam (Site 1) and 62 from Kampung Berua (Site 2) (Figure 1). The age and gender distribution are shown in Figure 1. Of those for whom age was accurately recorded, 49% were female and 45% male. There were notable challenges in the recruitment of children under five years old (accounting for only 11.5% of the total population sampled). All those with missing age data were adults.

**Figure 1.**
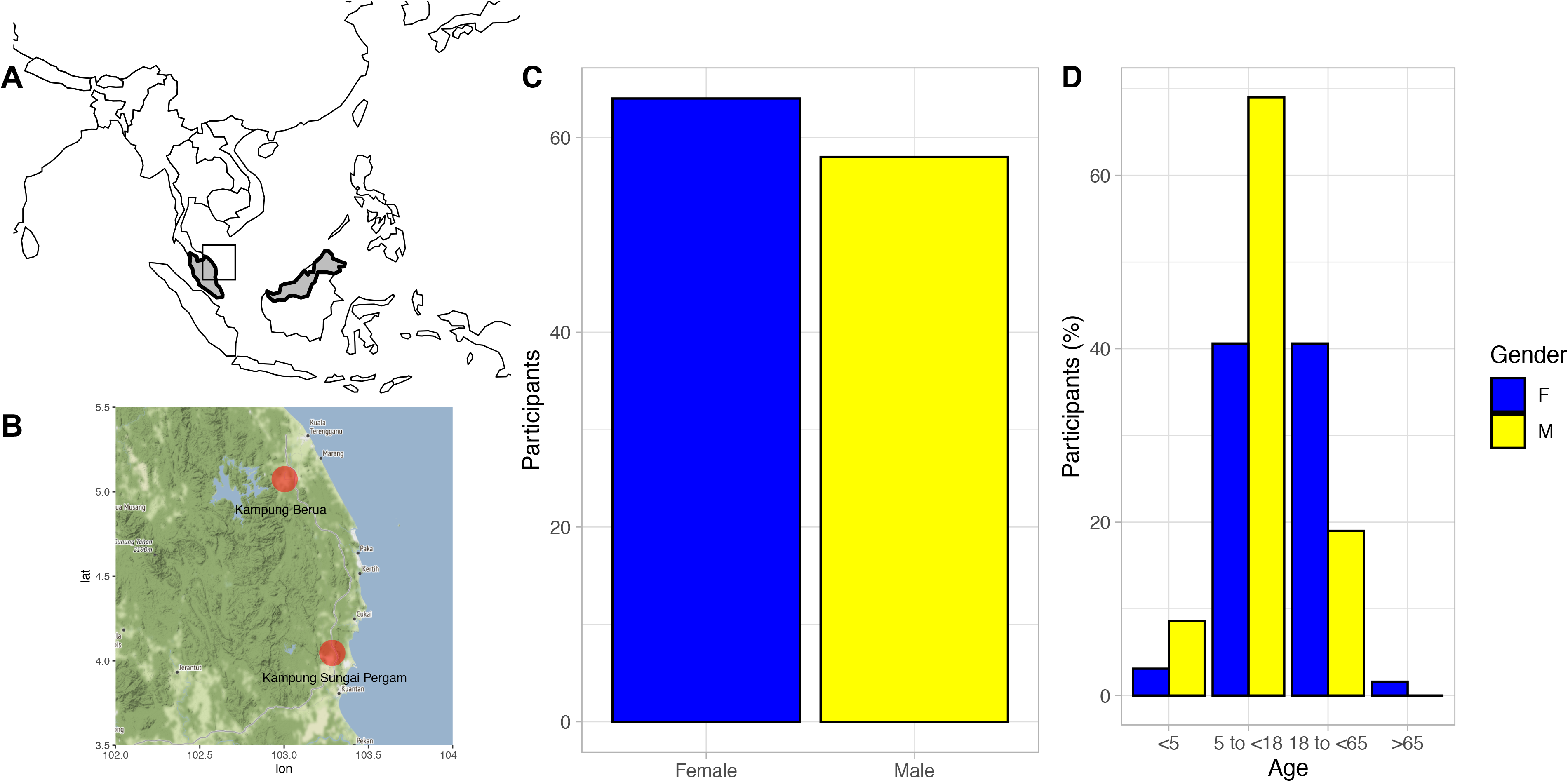
Location of Orang asli sites with age and gender demographics. The location of Orang Asli settlements were in the north-east of Peninsula Malaysia (A) to the south of the Terengganu state’s capital city, Kuala Terengganu (B). An even gender split was achieved (C) but with disproportionate recruitment in older children >5 years of age and adults less than <65 (D).

The carriage prevalence of the four most commonly isolated pathobionts in the anterior nares and nasopharynx are shown in Figure 2. Here carriage was defined as the isolation of a pathobiont from either of the two nasal swabs and for visual clarity all adults were combined into one age group (18-65). The most commonly isolated pathobionts were *S. aureus* and *S. pneumoniae* both with a total of 39 individuals colonised. The highest carriage for *S. aureus* was in the 18-65 age group at 47.2% (n=17/36; 95%CI: 30.9-63.5), followed by 30% (n=18/60, 95%CI: 18.4-41.6) in the 5 to 17 year olds. In contrast, the highest carriage of pneumococci was in the <5 and 5 to 17 year olds, with 57.1% (4/7) and 49.2% (30/61) respectively. Looking at the latter cases in more detail shows that the average age of a pneumococcal carrier in this age group was ~9 years old (range: 6-13). The next most carried bacterium was *H. influenzae*, isolated from seventeen individuals, the majority of which (88.2%) were in the 5 to 17 years old age group. In contrast, *M. catarrhalis* was rarely isolated (n=6), as was *N. meningitidis* (2), *P. aeruginosa* (n=3), *K. pneumoniae* (n=6) and 0-haemolytic *Streptococci* (n=7). Carriage and co-carriage are summarised in Table 1. Carriage of only *S. aureus* or *S. pneumoniae* accounted for 31.5% of the profiles observed. A limited number of co-carriage incidents were found which included seven and five cases of pneumococci isolation with *H. influenzae* or *S. aureus* respectively.

**Figure 2:**
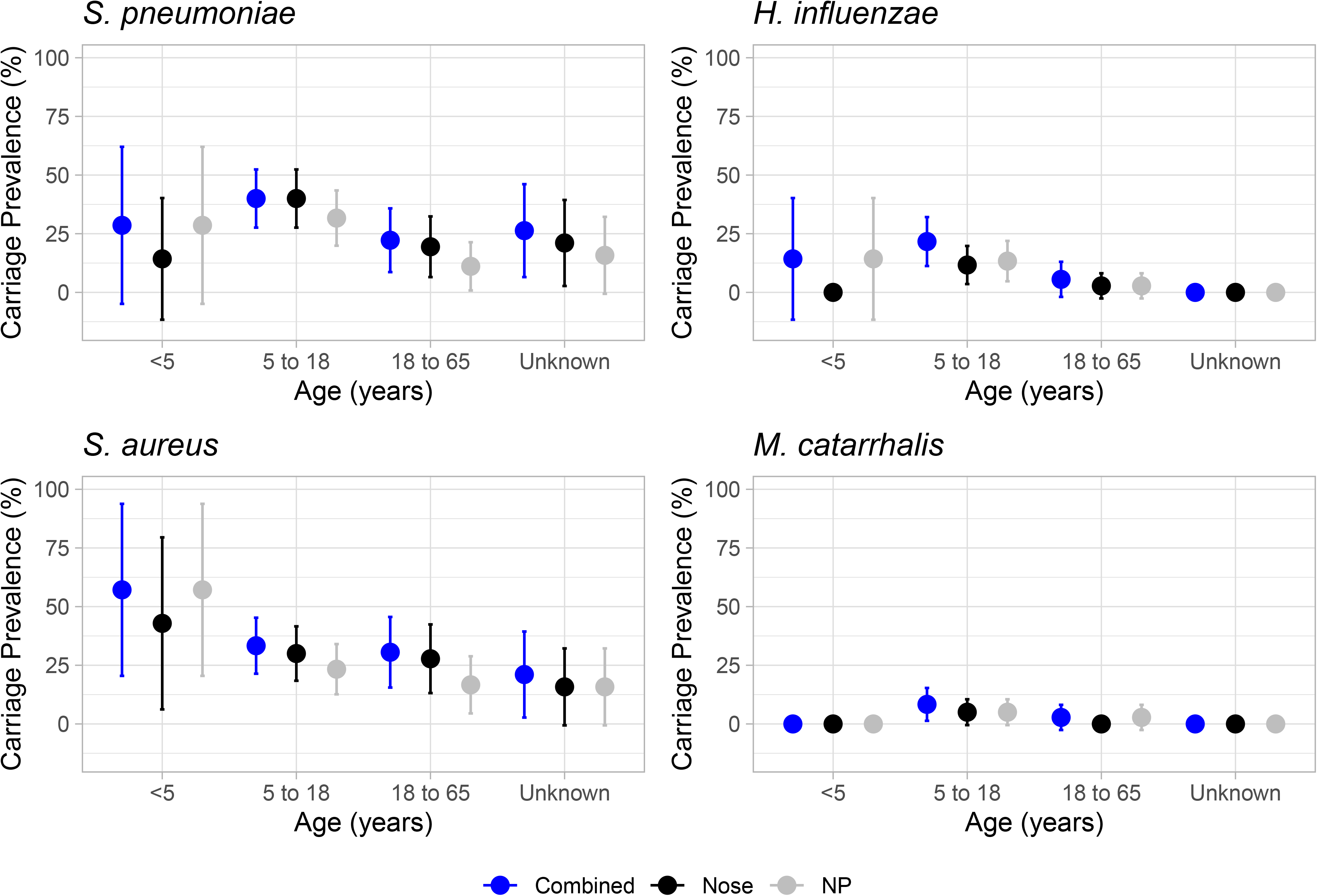
Carriage Prevalence of *S. pneumoniae, H. influenzae, S. aureus* and *M. catarrhalis*. Point estimates for percentage carriage prevalence of four key upper respiratory tract pathobionts are shown across three age groups <5, 5 to 17 and 18 to 65. Those aged 65+ are not shown due to limited numbers. All ‘Unknowns” were of adult age. Carriage was estimated overall i.e. a positive swab culture from either nasal or nasopharyngeal swab (dark blue), and also separately for each swab type (nasal - black; NP - grey). Error bars show 95% CI intervals.

**Table 1:**
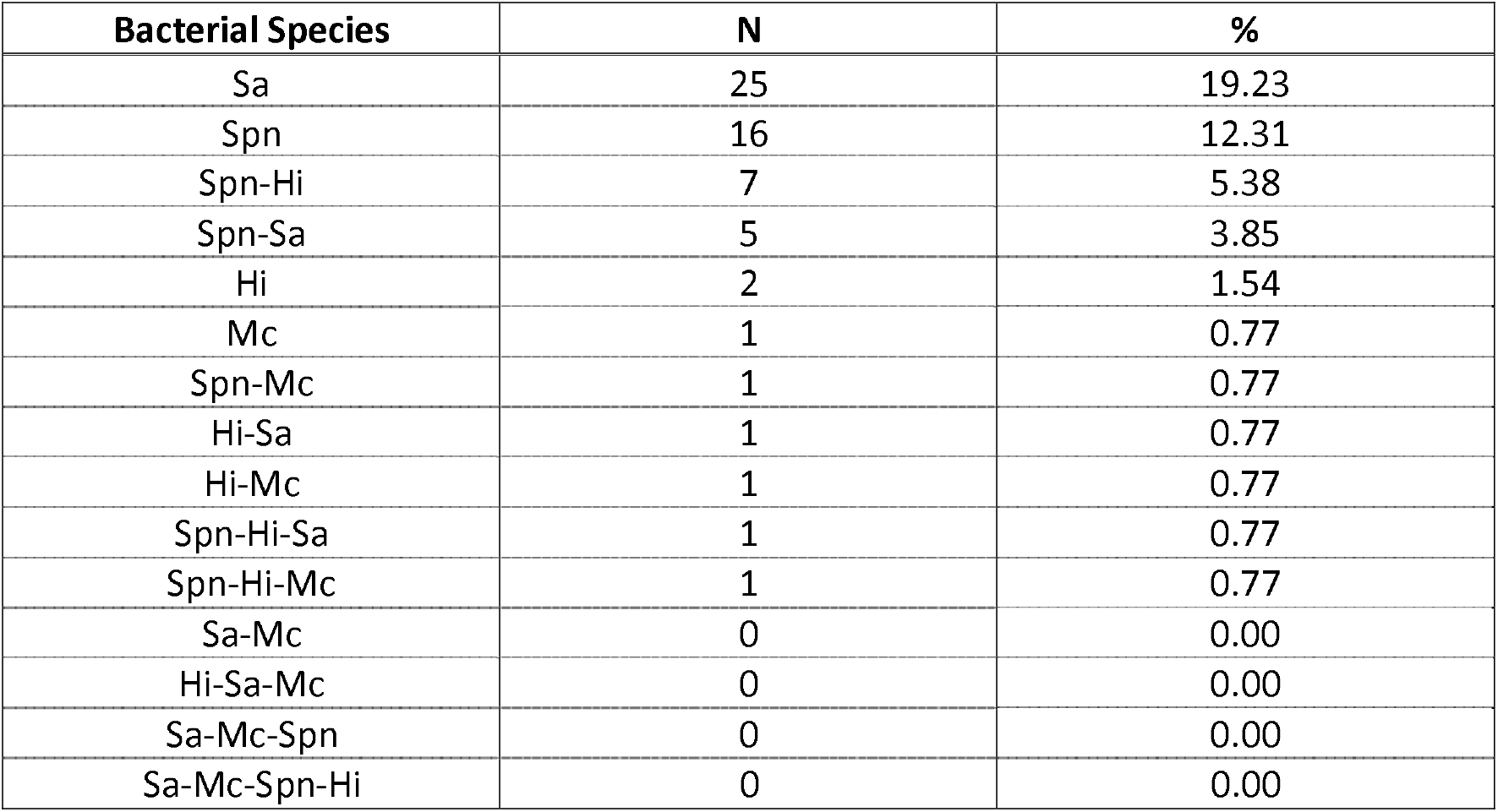
Multi-versus single-species carriage of *Streptococcus pneumoniae* (Spn), *Staphylococcus aureus* (Sa), *Haemophilus influenzae* (Hi) and *Moraxella catarrhalis* (Mc). Forty-seven individuals were culture negative. Seven had single-species profiles involving *Klebsiella pneumoniae* (n=3), α-haemolytic *Streptococci* (n=2) or *Pseudomonas aeruginosa* (n=2). A final thirteen had different multi-species profiles to those listed below.

Serotyping of pneumococcal isolates revealed 16 different serotypes (Figure 3). To avoid repeated sampling bias, if isolates recovered from nose and nasopharyngeal swabs in a single individual were the same serotype this was only counted once. The most common serotypes were the non-VT 23A (14.6%) and the VT 6B (9.8%). Overall, five of the 16 pneumococcal serotypes would be covered by Prevenar 7® (Pfizer, PCV7) (4, 6B, 14, 19F and 23F) with two additional serotypes covered by Prevenar 13® (Pfizer, PCV13) (3 and 6A). Only four of the serotypes (4, 14, 19F and 23F) are included in Synflorix® (GSK, PCV10). Six isolates were not serotypeable. All but one of the vaccine-type pneumococci (a single 23F) were isolated from the site at Kampung Berua (Site 2).

**Figure 3:**
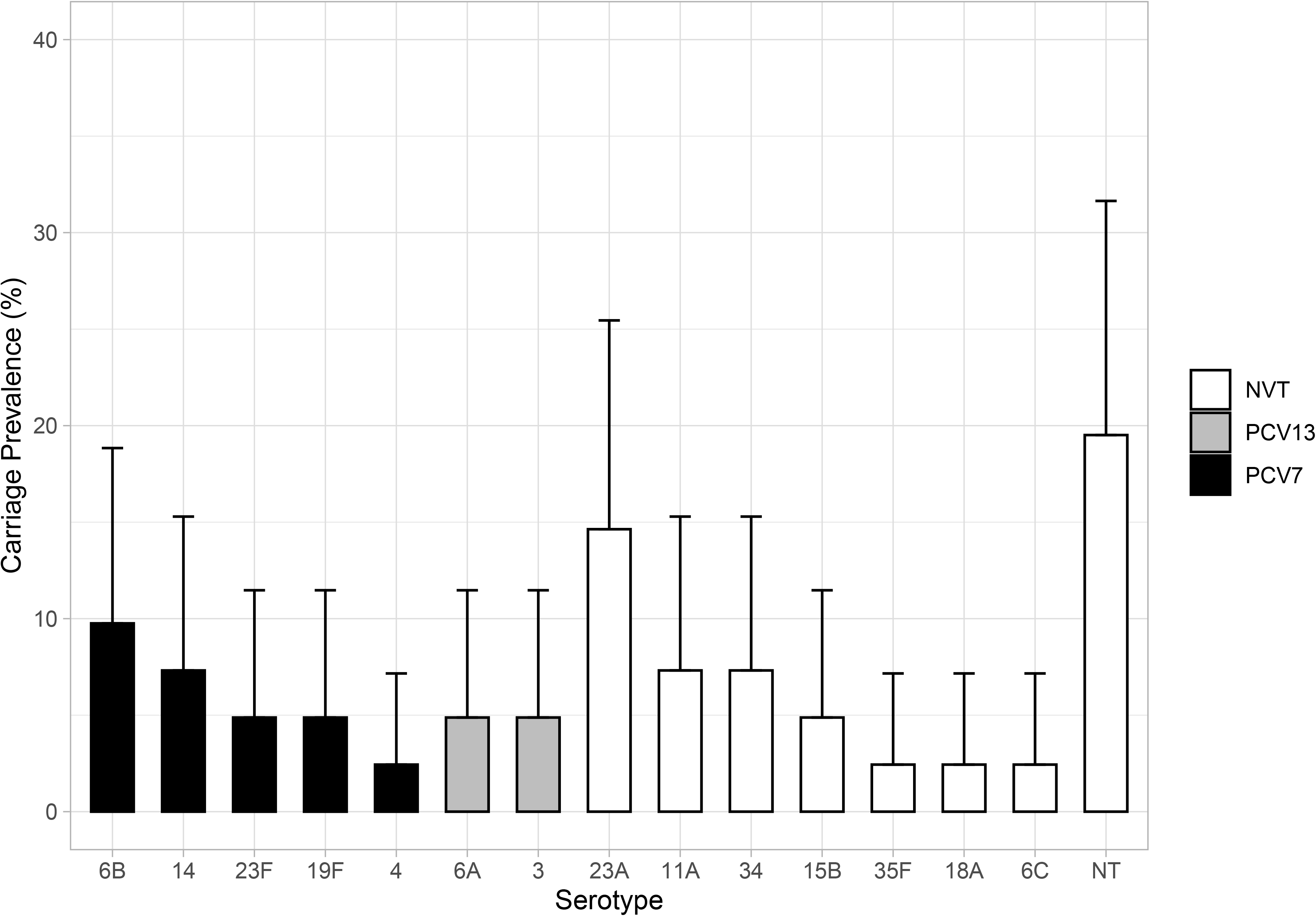
Prevalence of PCV7, PCV13 and non-vaccine pneumococcal serotypes. Bar plot with 95% CI showing prevalence of serotypes that are included in PCV7 (black), additional types covered by PCV13 (grey) or non-vaccine serotypes (white).

The extent of antimicrobial resistance for the key pathobionts is shown in Figure 4. Notable levels of resistance were observed for *S. aureus* where 73.3% (n=44) of isolates were resistant to penicillin, 33.0% (n=20) to tetracycline and 18.3% (n=11) to ciprofloxacin. However, all isolates were sensitive to cefoxitin, and all but one to chloramphenicol. Three of the *H. influenzae* isolates (15%) were resistant to penicillin, all of which were negative for β-lactamase activity. Of the *S. pneumoniae*, 17.9% (n=12) were resistant to tetracycline. No resistance was seen for *M. catarrhalis* against any antibiotic tested. *A. baumannii* were sensitive to meropenem and ciprofloxacin (n=14). Similarly, all *K. pneumoniae* were sensitive to all antibiotics tested including ceftazidime and meropenem.

**Figure 4:**
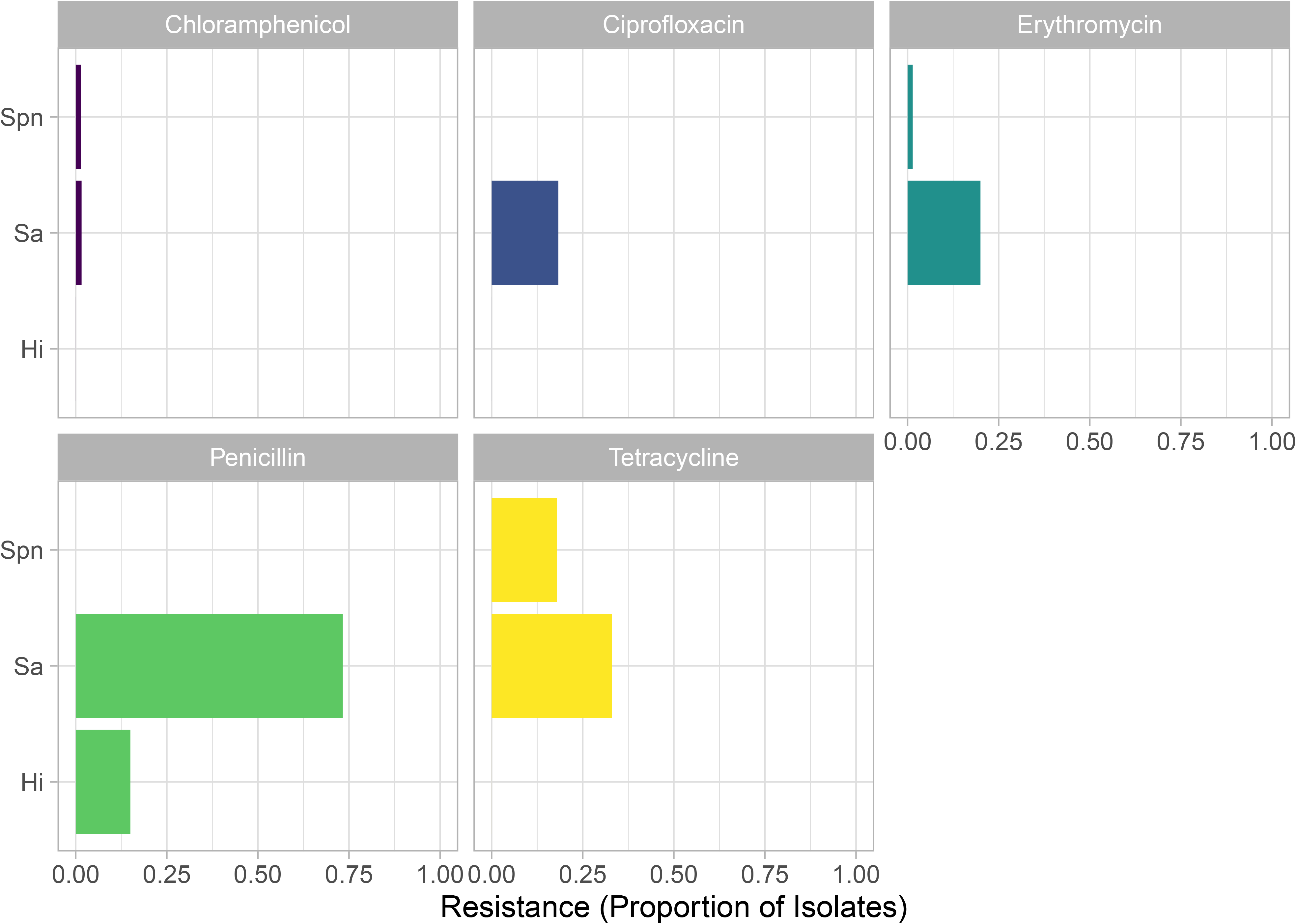
Antibiotic resistance of *S. pneumoniae* (Spn), *S. aureus* (Sa) and *H. influenzae* (Hi) Columns represent the proportion of isolates that were resistant to tetracycline, chloramphenicol, erythromycin, ciprofloxacin and penicillin.

The microbiome of the nasal (anterior nares) and oral (whole mouth) was examined using 16S rRNA sequencing of the V4 hypervariable region. Here, the median number of sequences per sample, including negative controls, was 11,431 with a maximum of 1,072,721. Eighteen nasal and two oral samples were excluded based on low numbers of ASVs (<1000). Alpha diversity (Figure 5A and 5B) was similar between all ages apart from nasal samples from those aged 5-17 and 50-65 where a significantly greater diversity was observed in the younger group (*p* = 0.0023); although notably fewer samples were available from the older participants. No clustering was observed using DPCoA for oral samples (Figure 5C - right column). In contrast, groups are clearly visible for nasal samples (Figure 5C - left column), with stratification based on age. PERMANOVA analysis of the Bray-Curtis distance showed that this was a significant difference (p = 0.001). Examining the most abundant phyla (Figure 6) showed nasal samples were dominated, as expected, by Firmicutes, Proteobacteria and Actinobacteria. For oral samples Bacteroidetes was a much more prevalent phyla in addition to Fusobacteria, although those same three phyla which dominated the nasal samples were also present at high levels here as well. Hierarchical clustering of Bray-Curtis dissimilarity scores revealed six clusters that could be characterised by the dominance of only one or two Genera (Figure 7). The largest cluster (n=28; 35.4%) contained individuals with a marked abundance of *Corynebacterium*, accounting for an average ASV relative abundance of 73.2% (range: 59.5 - 87.2%). The second largest cluster (n=16; 20.3%) also had a substantial proportion of *Corynebacterium*, in this case with *Dolosigranulum* as significant co-carried genus. Here, on average, *Corynebacterium* accounted for 48.88% and *Dolosigranulum* 31.6% of the relative abundance; combined these Genera accounted for between 71.5 and 89.4% of the ASV relative abundance of this profile. Two groups were then definable by the presence of *Moraxella*, one with and the other without *Haemophilus*. In the *Moraxella-dominated* profile, between 48.2 and 75.2% of the ASV relative abundance could be attributed to this Genus. When combined with Haemophilus this proportion dropped slightly from an average of 58.1% to 41.5% and here *Haemophilus* accounted for 13.2 to 47.9% of the profile. A small grouping of *Delftia/ Ochrobactum* profiles were also found. The final cluster consisted of only two profiles, where in one *Streptococcus* accounted for 63.8% and the second with *Streptococcus/Haemophilus* at 25.9 and 52.6% respectively. Interestingly, no *Staphylococcus* profiles were observed and none that were characterised by *Haemophilus* alone. No correlation between profile (in terms of genus composition) and culture outcomes for *S. aureus* or *S. pneumoniae* were seen (Figure 7). To determine if the abundance of either genus was related to culture positivity, the relative abundance of each was compared between samples from which either *S. aureus* or *S. pneumoniae* was isolated (Supplementary Figure 1). The relative abundance of Staphylococcal ASVs was not significantly different between groups, however those from whom *S. pneumoniae* were cultured did have a significantly higher relative abundance of Streptococci ASVs (*p* = 0.016); although this may be due to the presence of two profiles characterised by a significant dominance of *Streptococcus*. To explore the differences in microbiomes between the younger and older age groups, differentially abundant ASVs were identified using DESeq2 (Figure 8). The 5-17 year age group had ASVs classified as *Haemophilus* (including *H. influenzae), Moraxella* and *Streptococcus*. In contrast, *Propionibacterium, Peptoniphilus* and *Corynebacterium* ASVs were significantly increased in adults.

**Figure 5:**
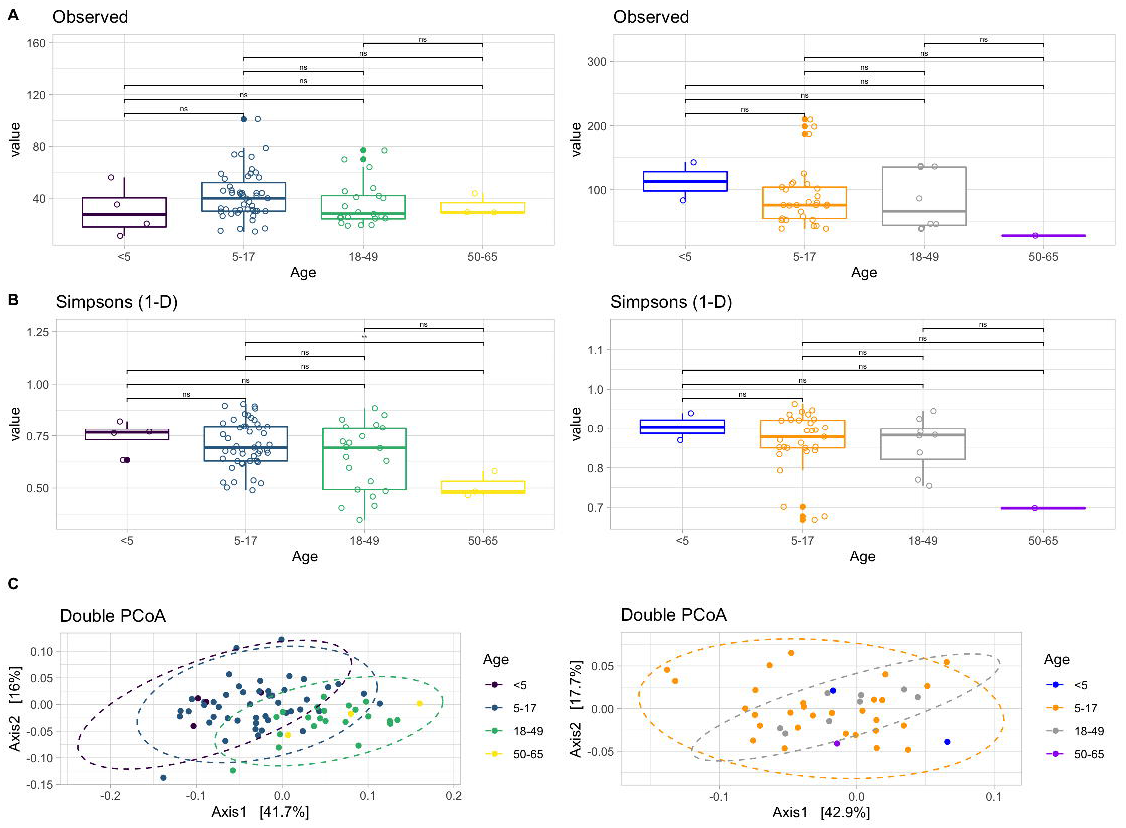
Alpha and Beta Diversity of Nasal (left column) and Oral (right column) Samples. Observed richness and Simpsons 1-D (a measure of diversity) are shown in rows A and B respectively. Only a significant difference (**) was observed for nasal samples for those aged 5-17 when compared against 50-65 (*p* = 0.0023). Beta diversity is shown using Double Principle Coordinates Analysis (DPCoA) (row C). Here, clusters are observed for nasal samples (left) with grouping based on age-group. No clusters were observed for oral samples (right).

**Figure 6:**
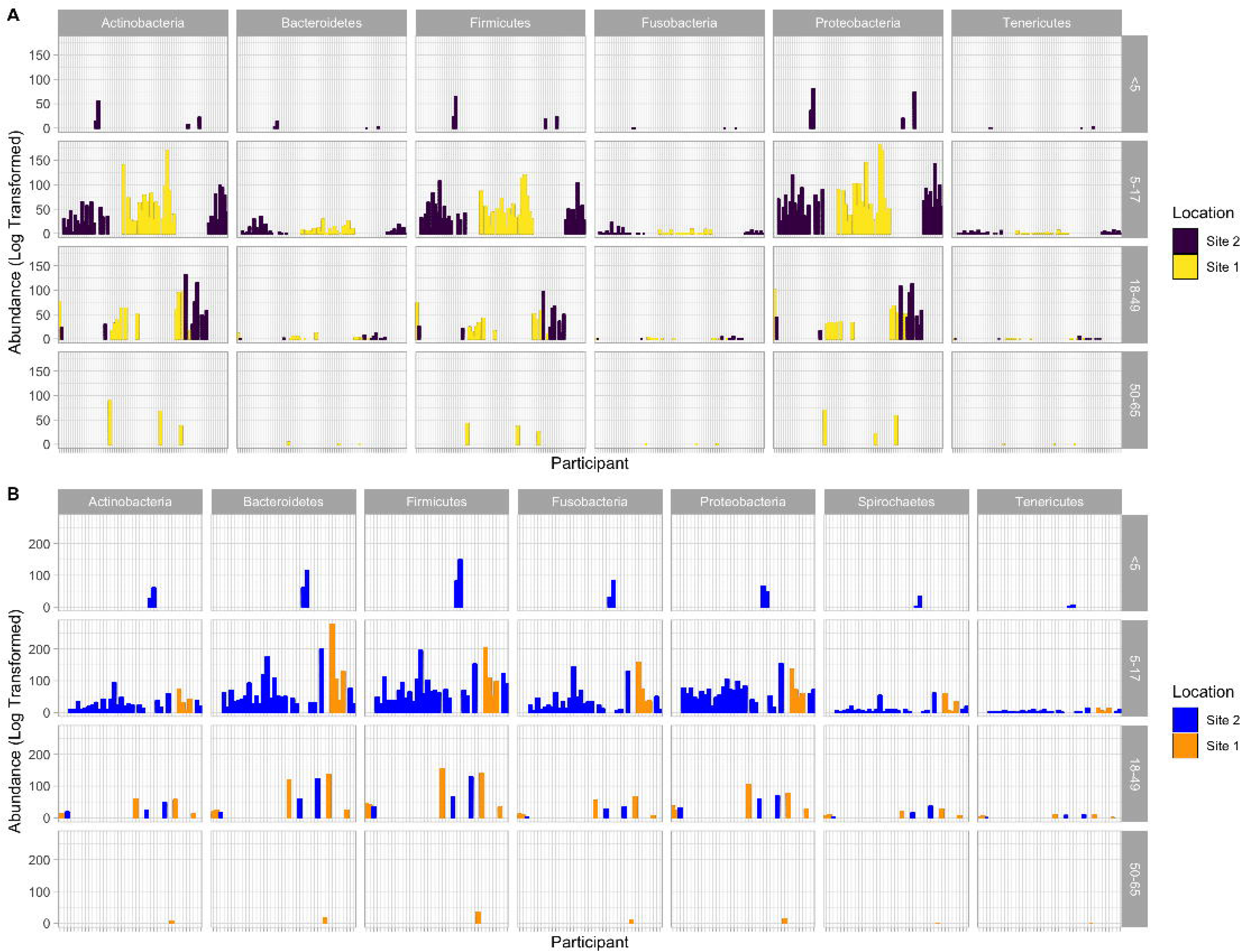
Log Transformed Abundances of Phylum-level ASV Classifications in Nasal (top) and Oral (bottom) Samples. Bars are coloured by site of location. Firmicutes, Actinobacteria and Proteobacteria were the most abundant phyla in Nasal samples with Bacteroidetes, Firmicutes and Proteobacteria the most common in Oral samples. A clear increase in Firmicutes in nasal samples from Site 2 (Kampung Berua) can be seen.

**Figure 7:**
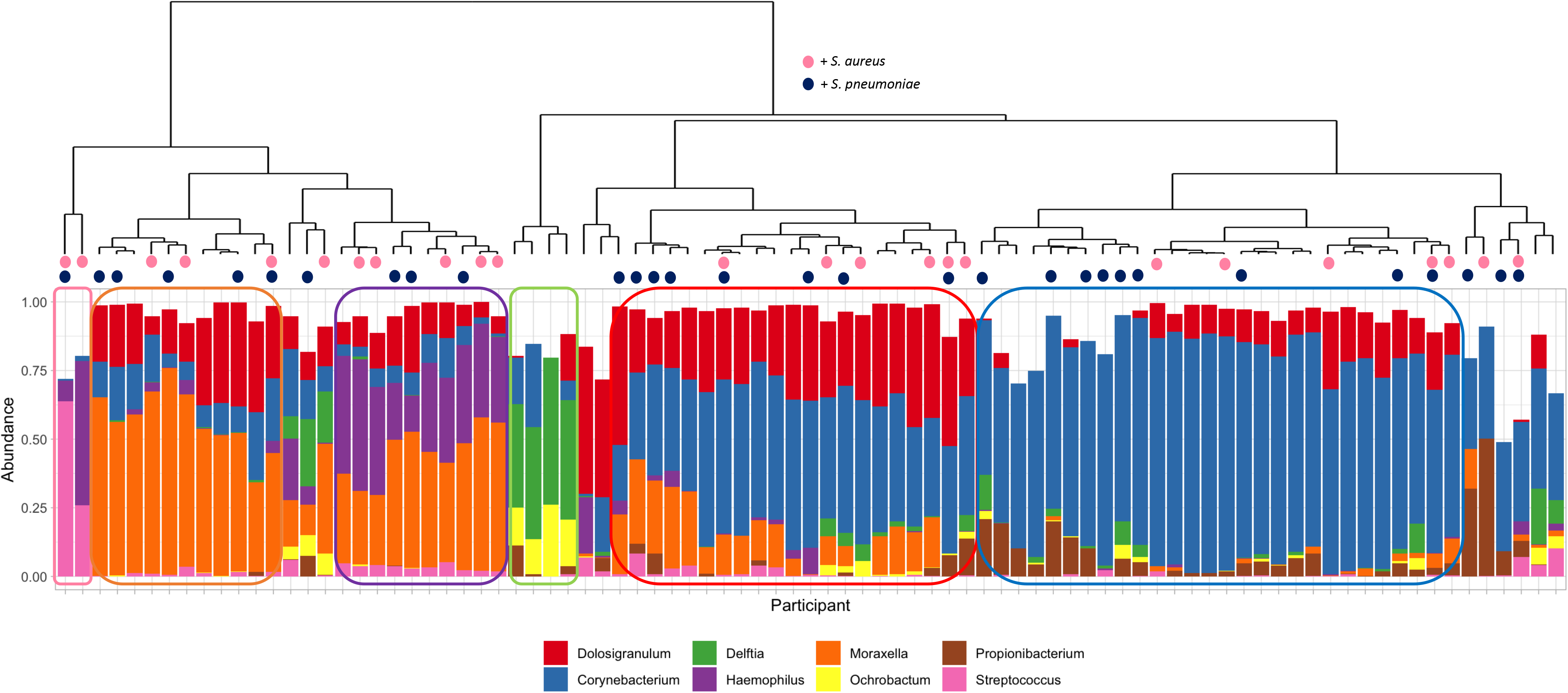
Hierarchical Clustering of Nasal Samples with Bray-Curtis Dissimilarity using Genus-level Relative Abundances. The dendrogram (top) shows clustering of samples with the below bar chart showing the relative abundance of the eight most commonly observed Genera. Six clear clusters were observed including a *Corynebacterium* dominant profile (n=28; 35.4%), a *Corynebacterium/Dolosigranulum* profile (n=16; 20.3%), a *Moraxella* profile (n=10; 12.7%) a *Moraxella/Haemophilus* profile (n=10; 12.7%), a *Delftia/ Orchobactum* profile (n=4; 5.1%) and a final group of two samples characterised by high *Streptococcus*, in one case with *Haemophilus*. Culture results for *S. aureus* (dark blue) and *S. pneumoniae* (pink) are shown as coloured circles at the tips of the dendrogram. There is no clear distribution of individuals who were culture positive for either bacterium.

**Figure 8:**
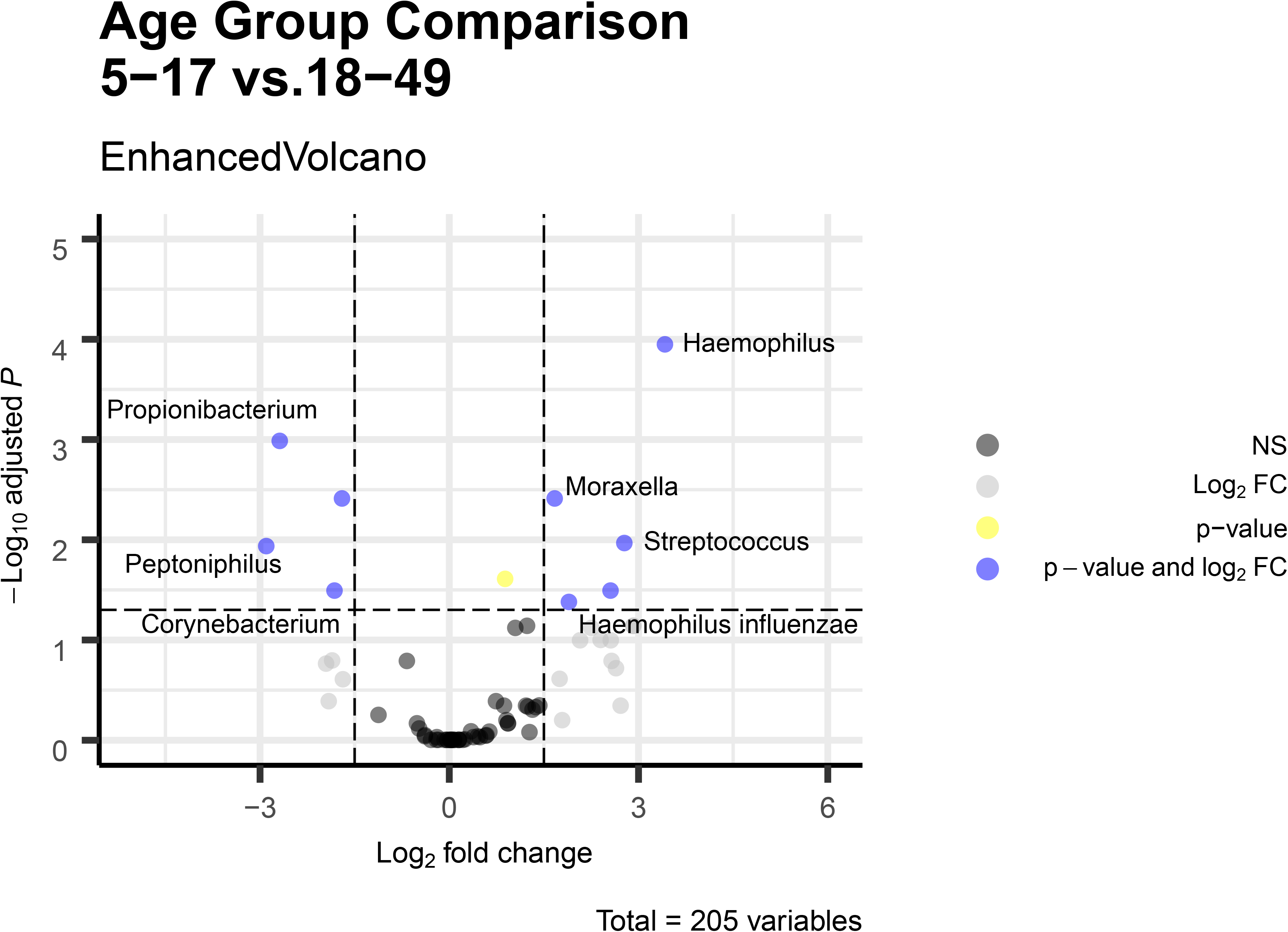
Differentially Abundant ASVs Between Older Children (5-17 years) and Adults (18-49) Volcano plot showing the ASVs that were differentially abundant in comparisons between the two age groups. Here a p-value cut-off of 0.05 and a Log_2_ fold change of 1.5 was applied. Those ASVs that pass both these thresholds are labelled where a genus and/or species taxonomic assignment was available.

*Streptococcus*, *Neisseria* and *Haemophilus* dominated the oral microbiota (Figure 9). Lower levels of *Prevotella, Rothia, Porphyromonas, Veillonella* and *Aggregatibacter* were also among the eight most observed genera. Streptococcal-dominated profiles were characterised by 43.7% (±15.7%) relative abundance of ASVs belonging to this genus. Those with a *Streptococcus/Haemophilus* profile still had 30.7% (±7.6%) Streptococci ASV relative abundance but with 15.0% (±10%) *Haemophilus*. The final profile, where clear dominance of one particular genus was observed, were those where *Neisseria* accounted for 30.6% (±12.5) of the relative abundance of all ASVs.

**Figure 9:**
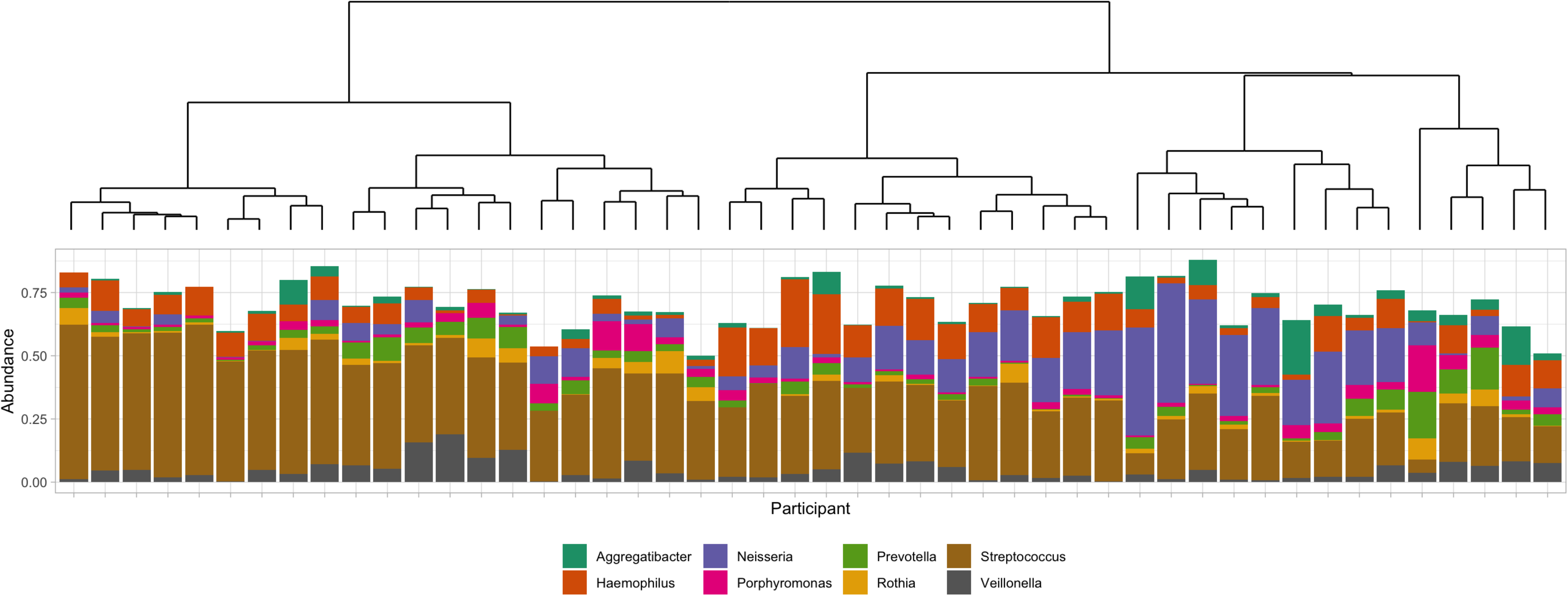
Hierarchical Clustering of Oral Samples with Bray-Curtis Dissimilarity using Genus-level Relative Abundances. The dendrogram (top) shows clustering of samples with the below bar chart showing the relative abundance of the six most commonly observed Genera. Three genera are seen to be responsible for the majority of ASVs - *Streptococcus* (brown), *Haemophilus* (orange) and *Neisseria* (purple). Profiles characterised by an overabundance of *Streptococcus* or *Neisseria* are seen, as too are those containing both *Streptococcus* and *Haemophilus*.

## Discussion

As a site that can be colonised by commensals, as well as opportunistic pathogens such as the pneumococcus [17], *S. aureus* [18] and *H. influenzae* [19], understanding the microbiology of the upper respiratory tract is key in tackling respiratory disease. The Anna Karenina principle of microbiomes, that “all healthy microbiomes are similar; each dysbiotic microbiome is dysbiotic in its own way” is a useful maxim for examining microbiomes in the context of disease [20]. However, from previous studies of communities that live unique, traditional lifestyles a clear challenge to the paradigm of health mirroring health has been shown [3, 8]. Here we examined the respiratory microbiomes of Orang Asli communities living traditional lifestyles in Terengganu, a rural state in the North-east of Peninsular Malaysia. These communities are known to face specific health challenges to that of the broader population of Malaysia, a country where respiratory disease is still a significant burden [21]. Our study showed a high prevalence of *S. aureus* carriage but interestingly not the presence of Staphylococci-dominated nasal microbiomes. Instead, *Corynebacterium*, with *Dolosigranulum* and *Moraxella* dominated as commensal flora. We also showed the potential impact that PCV introduction in Malaysia may have on circulating pneumococcal serotypes.

Although pneumococcal carriage can vary globally between 10 and 90% [22-24], the highest carriage prevalence is usually seen in young children, particularly those <5 years [24]. Although only a small number of this age group was sampled here (n=7), over half were colonised suggesting high levels of carriage. Given that in the 5-17 year age group the mean age was young at ~9 years old the high carriage (50%; 95%CI, 37.3 and 62.7%) seen in this age group supports this observation, being higher than has been seen elsewhere in older children [25] and similar to the 60.9% observed in Aboriginal children over 5 years old - a demographic known to experience high rates of invasive pneumococcal disease (IPD) [26].

As expected, the carriage in adults was low at 8.3% (95%CI, 0% to 17.4%), which is similar to the NP carriage prevalence of 11.1% (95%CI, 9.8 and 12.6%) that has been seen in the USA [27], but higher than recent surveys in the UK which found a 2.8% (95%CI: 1.2-5.5) prevalence in this age group [25]. As for serotypes, whilst there is growing epidemiological data for the region [28], there is limited data for Malaysia. A study of 245 cases of paediatric IPD between 2014 and 2017 showed that serotypes 14, 6B, 19A, 6A and 19F were the most common, accounting for ~75% of disease [29]. This is concordant with this study where serotypes 14 and 6B were also the most common in carriage with 6A and 19F being identified. Of the non-VT serotypes isolated only 34, 35F and 18A did not feature in those identified in the IPD study. The lack of 19A is intriguing but we suspect a consequence of the relatively low sampling. Both PCV10 and PCV13 are available in Malaysia however only in private practice and therefore the presence of VT serotypes is not unsurprising. These data do suggest introduction of PCV into the national immunisation programme as planned may prove efficacious.

Only one of the children less than 5 years old was culture positive for H *influenzae* but given the low numbers in this age group this must be interpreted with caution. Whilst certainly higher than in the UK where only 6.5% *H. influenzae* carriage was seen in older children [30], the 25% (95%CI, 14.0 and 40.0%) found here is similar to that observed in other developing countries such as Kenya [31], but lower than the 52.8% seen in Aboriginal children aged between 5 and 15 years old [26]. Whether this increased carriage translates into disease risk is difficult to determine; there is no data on the burden of otitis media among Orang Asli, although the country wide incidence in children <12 year olds is low at 2.3% for the Asia-Pacific region [32].

The prevalence of *M. catarrhalis* at 6.7% (95%CI, 0.35 and 13.0%) was very similar to that of our previous UK study where 5.7% of the same age group were colonised [30], a level also observed in other western countries [33]. This is a stark contrast to the 67% in Aboriginal children of a similar age [26], however this low level of carriage (6%) has been seen in Warao Amerindians in Venezuela [34] - arguably a similar demographic to the one being presented here.

*S. aureus* carriage is known to vary markedly, by age but also by geographic location [35]. Although the lowest prevalence observed here was in <5 year olds (28.6%; 95%CI: 0-62.0), which is surprising as one would expect the highest carriage would be seen in this age group, it could be either a feature of the low recruitment or that the average age of this group was 2.5 and carriage is known decline between 1 and 10 [18]. As for the carriage in adults, a previous study in Malaysia of 384 adults (students with an average age of 25) found a carriage prevalence of 23.4% [36] which is much lower than the 47.2% in our population of Orang Asli. A similarly high level of adult carriage (41.7% and 57.8% at two separate sampling timepoints) was observed in Wayampi Amerindians who, in terms of remote living, are similar to the Orang Asli [37]. Our own research has previously shown adult carriage at 24.4% in a UK population [30] and thus the differences we observed are unlikely to be methodological. More likely it is a consequence of greater co-habitation (the average household size of our communities being 5.2 people) and reduced access to hygiene, both of which have been highlighted as reasons why *S. aureus* carriage has declined in other parts of the world [18].

Of the ESKAPE pathogens isolated and tested only *S. aureus* exhibited any substantial level of resistance; importantly all isolates were sensitive to methicillin. Whilst there appears to be limited resistance it is important to note that this represents the only data on the AMR in these communities and is therefore merely a starting point for future studies. This is particularly important given that AMR burden in indigenous populations has been seen to increase elsewhere [38].

URT microbiome research has shown that the composition and/or dominance of particular bacterial taxa can be indicative of nasal community state types (CSTs). Previously, in a study of adults in the USA, seven CSTs were identified that were characterised by *S. aureus* (CST1), or other *Staphylococci* (CST3), a mix of Proteobacteria including *Escherichia* and *Enterobacteriaceae* sp. (CST2), *Corynebacterium* either with *Propionibacterium acnes* (CST4) or without (CST5), *Moraxella* (CST6) and *Dolosigranulum* (CST7) [39]. It is here that we see the most startling contrasts to out Orang Asli population. Although we also identified seven clusters based on what is/are the dominant taxa we did not see anything resembling CST 1 or 3 (*Staphylococci)*, and no CST2. The lack of a *Staphylococcus* dominated profiled is an enigma given the high carriage rate we observed. We hypothesise that the high prevalence of profiles similar to CST5, *Corynebacterium* dominated, along with another that is defined by high abundance of both *Corynebacterium* and *Dolosigranulum*, a composition that has been found in the nasopharynx [40], explains this absence and is based on previous work showing that *Corynebacterium* reduces *S. aureus* carriage [41]. A similar interaction may also explain the absence of *Streptococci* [42] - although we recognise in both cases that our hypothesis requires further testing due to the identification of both *S. aureus* and pneumococcal carriage among our participants. A *Moraxella* dominated profile similar to CST6, is also present in the Orang Asli, a profile which has also been observed in healthy adults in a European setting [43], although we also see a *Moraxella-Haemophilus* profile which hasn’t been described previously. The *Streptococcus* profiles are both from older children (6 and 10 years old) and this not uncommon as a profile among this age group [44]. The significant proportion of individuals who harbour profiles that are being considered to be potentially protective either in terms of acute respiratory infections [45] or chronic disease such as asthma [46-48], is intriguing. Lastly, whilst *Delftia* is not uncommon as an oral bacterium [49], given the common finding of *Delftia* and *Ochrobactum* as common contaminants in microbiome research [50] we interpret these four samples with extreme caution. The presence of these profiles is in spite of our efforts to control for such contaminants through the collection and sequencing of control swabs. Epidemiologically, all four samples come from adult females, but two each from Kampung Berua and Kampung Sungai Pergam. The samples are below the 25% quantile for ASV number (n=3831). It is possible that these reflect true profiles, but it is impossible to rule out process error during sample handling, extraction and/or sequencing.

The microbiota recovered from the oral cavity was substantially more diverse than the nasal samples, as expected [49]. The top three genera were unremarkable in terms of what is considered to be most commonly observed, mirroring exactly that found in a study of 447 datasets deposited as part of large-scale microbiome projects [51]. A previous study of Temiar, Orang Asli in Perak found a significant difference in oral microbiota compositions between males and females [16], however a PERMANOVA analysis of the data presented here showed no such distinction (*p* = 0.41).

Whilst this piece of research was highly novel as a first glimpse into the respiratory microbiology of Orang Asli communities, it could nevertheless have been strengthened in a number of ways. Firstly, it was only possible to visit two sites on a single occasion, and to conduct a pragmatic survey of the communities. As a consequence, the study lacks follow-up and longitudinal data. Whilst we were very successful at recruiting older children and adults, future studies will have to identify how to recruit greater numbers of younger children in particular. In terms of pathobiont carriage, serotyping of more than one pneumococcal isolate from each individual would have enabled a determination of the prevalence of multiple serotype carriage. Serotyping of *H. influenzae* would also have been informative, although future work on exploring the genomics of all isolates will address this in more detail. This additional insight could also be expanded to include all the pathogens/pathobionts isolated. As with all microbiome research, longitudinal sampling taking into account an individual’s temporal variability, the seasonality and medical history would be desirable.

## Conclusions

Here we present the first study of Orang Asli airway microbiomes and pathobiont microbiology, including the antimicrobial resistance profiles of ESKAPE pathogens. Important findings include the prevalence of pneumococcal serotypes that would be covered by pneumococcal conjugate vaccines, supporting their introduction as part of a national immunisation programme. The high prevalence of *S. aureus* carriage is noteworthy and warrants further study. In terms of the airway microbiomes the dominance of *Corynebacterium*, additionally with *Dolosigranulum* and *Moraxella* in nasal profiles is particularly intriguing and future work should explore these commensals in the context of burden and susceptibility to both acute and chronic respiratory conditions in these communities.

## Methods

Study sites and participants: Two Orang Asli villages were visited in August 2017 – Kampung Sungai Pergam in Kemaman district and Kampung Berua in Hulu Terengganu district. Both sites are located in the state of Terengganu which lies in the north-east of Peninsular Malaysia. Participant recruitment was with consent, across all ages with no exclusion criteria. Questionnaires were used to capture participant metadata which included gender, age, number of dwelling co-occupants and occupation in addition to health-related questions including current or recent (within the last month and last three months) respiratory symptoms, antibiotic use and vaccination status. Questionnaires were translated into Malay by A.S.M.H., A.R., M.A.R., and C.C.Y., and conducted by research assistants from the Faculty of Applied Social Sciences, Universiti Sultan Zainal Abidin, who were trained on how to administer the questionnaires for the purpose of this study.

Swab collection: A total of four swabs were taken from each participant. A nasal swab (anterior nares) and a nasopharyngeal swab, using rayon tipped transport swabs containing Amies media with charcoal (Medical Wire and Equipment, Corsham, UK), were used for bacterial pathobiont isolation. A further nasal swab, taken from the un-swabbed nostril, and an oral (whole mouth) swab, supplied by uBiome Inc. (San Francisco, USA), were taken for microbiome analysis.

Bacteriology: Isolation of the common respiratory pathobionts *S. pneumoniae, H. influenzae, M. catarrhalis, N. meningitidis, S. aureus, K. pneumoniae* and *P. aeruginosa* was done by culture. Swabs were plated onto CBA (Columbia blood agar with horse blood), CHOC (Columbia blood agar with chocolated horse blood), CNA (Columbia Blood Agar with Colisitin and Naladixic Acid), BACH (Columbia Agar with Chocolated Horse Blood and Bacitracin), GC (Lysed GC Selective Agar) and, for *P. aeruginosa*, CFC (*Pseudomonas*CFC Selective agar) (all Oxoid, UK). Primary identification of all bacterial species was by colonial morphology. Swabs were then vortexed in skim milk, tryptone, glucose and glycerine (STGG), and 10jiL of the suspension was then Serotyping of S. *pneumoniae*. Pneumococcal isolates were serotyped by slide agglutination reactions using a Neufeld *S. pneumoniae* antisera kit following manufacturer’s guidelines (Statens Serum Institute, Copenhagen, Denmark).

Antimicrobial Resistance Testing: All bacteria were phenotypically tested for antibiotic resistance using antibiotic discs and/or minimum inhibitory concentration (MIC) strips, in accordance with EUCAST. Firstly, 10μL (a suspension of cells in liquid STGG) of each isolate was plated onto CBA (Oxoid, UK) or CHOC agar (Oxoid, UK). *M. catarrhalis, S. pneumoniae, S. aureus* and *K. pneumoniae* isolates were plated on CBA, whilst *H. influenzae* and *N. meningitidis* isolates were plated onto CHOC agar. Plates were incubated for 24 hours at 37°C in 5% CO_2_. Pure colonies were added to 1ml of saline to get an inoculum of 0.5 McFarland. For *M. catarrhalis, S. pneumoniae, H. influenzae* and *N. meningitidis*, a sterile swab was used to spread this inoculum over Mueller-Hinton agar + 5% defibrinated horse blood and 20 mg^-1^ β-NAD plates (MHF, Oxoid, UK). For *S. aureus* and *K pneumoniae* a sterile swab was used to spread the inoculum over Mueller-Hinton agar plates (MH, Oxoid, UK). Antibiotic discs (Oxoid, UK) (four per plate) or MIC strips (E-tests; Oxoid, UK) (one per plate) were added and plates were incubated at 37°C in 5% CO_2_ for 18 hours (±2 hours). *M. catarrhalis* were tested with amoxicillin-clavulanic acid (2-1μg), cefotaxime (5μg), ceftriaxone (30μg), erythromycin (15μg), tetracycline (30μg), chloramphenicol (30μg), ciprofloxacin (5μg) and meropenem (10μg) antibiotic discs. *S. pneumoniae* were tested with oxacillin (1μg), erythromycin (15μg), tetracycline (30μg) and chloramphenicol (30μg) antibiotic discs. *H. influenzae* were tested with benzylpenicillin (1μg), tetracycline (30μg), chloramphenicol (30μg) and ciprofloxacin (5μg) antibiotic discs as well as erythromycin (15μg) MIC strips. *S. aureus* were tested with benzylpenicillin (1μg), erythromycin (15μg), cefoxitin (30μg), tetracycline (30μg), chloramphenicol (30μg) and ciprofloxacin (5μg) antibiotic discs. *K. pneumoniae* were tested with amoxicillin - clavulanic acid (20-10μg), cefotaxime (5μg), ciprofloxacin (5μg), meropenem (10μg) and ceftazidime (10μg) antibiotic discs. *N. meningitidis* were tested with amoxicillin, benzylpenicillin (1|ig), cefotaxime (5μg), ceftriaxone (30μg), chloramphenicol (30μg), ciprofloxacin (5μg) and meropenem (10μg) MIC strips.

16S rRNA Sequencing: The V4 region was amplified using primers 515F (5’-GTGCCAGCMGCCGCGGTAA-3’) and 806R (5’-GGACTACHVGGGTWTCTAAT-3’) to generate an amplicon of 460 bp [52] at uBiome Inc. (San Francisco, USA). Samples were individually barcoded and sequenced on a NextSeq 500 (Illumina, San Diego, USA) to generate 2 × 150 bp paired-end reads.

Microbiome Analysis: Initial data handling was done using QIIME 2 2018.8 [53]. Only forward reads were processed owing to low quality scores of reverse reads. Raw sequence data were denoised with DADA2 [54]. Amplicon sequence variants (ASVs) were aligned with mafft [55] (via q2 J alignment) and a phylogeny constructed using fasttree2 [56]. Taxonomic classification was done using the q2 feature classifier [57] classify Jsklearn naïve Bayes taxonomy classifier using the Greengenes 13_8 (99%) reference sequences [58]. The feature table, taxonomy, phylogenetic tree and sample metadata were then combined into a Phyloseq object using qiime2R [59] via qza_to_phyloseq. All further analysis was done in R v3.6.0 [60] in RStudio and figures were produced using the package ggplot2 [61]. Phyloseq v1.29.0 [62] was used following a workflow previously described [63]. Potential contaminants were identified by prevalence in the blank swab controls and removed using the R package ‘decontam’ [64]. Samples with <1000 ASVs were excluded and taxa present in less than 5% of samples were removed using the prune_taxa() phyloseq function. Alpha diversity was calculated using the estimate_richness(). The R function stat_compare_means() was used to compare age groups using the nonparametric Wilcoxon test. Beta diversity was determined using Double Principle Co-ordinates Analysis and plot_ordination(). Hierarchical clustering was done using Bray-Curtis Dissimilarity calculated using vegist() from the R package vegan [65]. Differentially abundant ASVs were identified using the DESeq2 [66] package using an adjusted p-value cut-off of 0.05 and a Log_2_ fold change of 1.5.

## Data Availability

Sequence files and associated metadata have been deposited in the European Nucleotide Archive (ENA) in project PRJEB38610 under experiment accessions ERX4147052 to ERX4147288.

**Abbreviations**
AMR: antimicrobial resistance
ASV: amplicon sequencing variant
PCV: pneumococcal conjugate vaccine
VT: vaccine type pneumococci
NVT: non-vaccine type pneumococci
ESKAPE: *Enterococcus faecium, Staphylococcus aureus, Klebsiella pneumoniae, Acinetobacter baumannii, Pseudomonas aeruginosa*, and *Enterobacter* spp.

## Ethics Approval and Consent to Participate

Ethical approval for this study was provided by Universiti Sultan Zainal Abidin (UniSZA) Ethics Committee: approval no. UniSZA/C/1/UHREC/628-1(85) dated 27th June 2016, the Department of Orang Asli Affairs and Development (JAKOA): approval no. JAK0A/PP.30.052Jld11 (42), and by the University of Southampton Faculty of Medicine Ethics Committee (Submission ID: 20831). Informed consent was taken for all participants, with parents/guardians providing consent for those <17 years old. Participation in the study involved reading (or being read) and understanding the translated participant information sheet, and the completion of a consent from and questionnaire. This process was facilitated by native speakers.

## Consent for Publication

Not applicable.

## Authors Contributions

DWC and SCC conceived the project with YCC. DE, RAA and HM were the UoS contingent who undertook the sampling and helped with study planning and design. AGA, NIAR, SI, MSR, RMA, AAA, NKE, SA, CHC MHAS and RA helped with study design, coordinated sampling visits to Orang Asli settlements, liaised with those communities and assisted with sampling and questionnaires. AGA did the microbiological culture and antibiotic testing with RAA and DE. DWC carried out the data analysis and wrote the paper. All authors provided critical feedback and helped shape the manuscript.

## Acknowledgments

We would like to thank the Department of Orang Asli Affairs and Development (JAKOA), Additionally, the authors would especially like to thank the communities of Kampung Sungai Pergam and Kampung Berua for their support and participation in this study.

---

**Supplementary Figure 1: Comparison of the relative abundance of Streptococcal and Staphylococcus ASVs between those that were culture positive or negative for *Streptococcus pneumoniae* or *Staphylococcus aureus. P* values were calculated using the nonparametric Wilcoxon test.**

